# Benchmarking RNA-seq Tools for Real-World Diagnostic Applications

**DOI:** 10.64898/2026.01.27.26344940

**Authors:** Sarah Silverstein, Kaushik R Ganapathy, Sandra Donkervoort, Véronique Bolduc, Ying Hu, Justin Moy, Prech Uapinyoying, Svetlana Gorokhova, Vijay Ganesh, Ben Weisburd, Rotem Orbach, A Reghan Foley, Pejman Mohammadi, David R Adams, Carsten G Bönnemann

## Abstract

**Background:** Pediatric neuromuscular diseases are genetically and clinically heterogeneous. A substantial proportion remain without a definitive genetic diagnosis despite available clinical molecular testing. RNA-sequencing (RNA-seq) can be used to complement genome or exome sequencing to elucidate or to identify the functional impact of variants of uncertain significance, but when manually analyzed is limited to candidate DNA variants or phenotype-driven gene lists. Open-source computational tools have been developed to systematically and unbiasedly analyze RNA-seq data for aberrant splicing, expression, or allelic imbalance. However, best use practice of these tools is yet to be established.

**Methods:** To assess the performance of selected tools, we collected RNA-seq from 97 previously diagnosed samples to establish a truth set for benchmarking. Pathogenic variants were categorized as: true positives with confirmed aberrant RNA events and true negatives with no transcriptomic effect. We assessed performance of eight commonly used tools for splicing, expression and allelic imbalance analysis. We then applied the optimal strategy to 74 undiagnosed RNA-seq samples to identify new candidate diagnoses.

**Results:** Across 68 diagnosed probands with aberrant RNA events, tools correctly identified 28 diagnoses. Splicing analysis tools provided most of the findings, but allelic imbalance tools uniquely identified 4, underscoring their value. Conversely, the false positive rate was highest for the splice tools and lowest for expression analysis. Application of tools led to identification of candidate variants for only 9 out of 74 undiagnosed patients.

**Conclusions:** Inclusion of RNA-seq tools can expedite variant prioritization, characterization and interpretation in the diagnostic pipeline but remain complementary to manual analysis of loci where candidate variants were identified by DNA sequencing.

## Background

Pediatric neuromuscular disorders are characterized by broad genetic and clinical heterogeneity, with approximately 30–60% of cases remaining molecularly undiagnosed following exome or genome sequencing[1–4]. Recent reports suggest that incorporating RNA-seq data derived from clinically accessible tissues such as whole blood, peripheral blood mononuclear cells, fibroblasts, urine, or muscle can increase diagnostic yield by an additional 2.7–32%[5–17]. In a typical clinical-diagnostic bioinformatics workflow of DNA sequencing, variants are prioritized based on phenotypically-driven analysis, possibly identifying candidate variants with effect on splicing or gene/protein expression. If RNA-seq data is obtained, splicing and expression of the genes with these variants are then manually inspected in the RNA-seq BAM file data. This process is time-consuming and, despite the availability of published guidelines, remains heavily reliant on the expertise of individual analysts and the availability of appropriate control and tissue specific datasets[18].

Recently, open-source computational tools have been developed to automate the analysis of transcriptomic data in the context of rare disease[19–24]. These tools are designed to identify aberrant RNA events—such as abnormal splicing, aberrant gene expression, or allelic imbalance—in a systematic and agnostic manner. Unlike manual methods, these tools employ an RNA-first strategy, enabling transcriptome-wide analysis independent of prior DNA variant prioritization. This approach has the potential to identify pathogenic events that may be overlooked during manual review. Applying open-source strategies to RNA-seq datasets derived from clinically accessible tissues has resulted in modest diagnostic yield, with the highest yield in cases where initial DNA sequencing identified a variant of uncertain significance (VUS) predicted to alter splicing and prioritized [5–7,15,17,25–28]. In cases without any diagnostic leads, RNA-first strategies have resulted in limited diagnostic yield [5–7,15,17,25–27]. To date, comprehensive benchmarking of these tools using previously diagnosed samples harboring pathogenic variants leading to known aberrant RNA events has yet to be undertaken. Furthermore, prior studies have typically sequenced large batches of samples and analyzed them collectively to mitigate batch effects. This strategy does not reflect real-world clinical constraints, where disease-relevant tissue is often limited, requiring evaluation of open-source tools using diverse approaches.

We assessed a range of RNA-seq analysis tools to identify those best suited for our diagnostic pipeline and to assess their limitations. The most effective strategy was then applied to a set of clinically well-characterized, undiagnosed cases to enable candidate variant discovery. We also evaluated the capacity of RNA-seq to function as a standalone diagnostic tool, versus being utilized as an adjunct to DNA-based approaches.

## Methods

### Patient Recruitment

Patients were referred by their neurologist or geneticist. Written informed consent and age-appropriate assent for study procedures were obtained by a qualified investigator [protocol 12-N-0095 approved by the National Institute of Neurological Disorders and Stroke, National Institutes of Health IRB]. Medical history was obtained, including clinical evaluations and muscle biopsy, were performed as part of the standard diagnostic examination. Samples for research-based testing, including skin, blood and muscle biopsy samples, were obtained using standard procedures. Patients with confirmed genetic diagnosis were included as true positive or true negative benchmarking samples, and patients without candidates after evaluation were included for our undiagnosed samples analysis (Supplemental Methods).

### RNA-seq Alignment

Quality control of FASTQ files were assessed with FastQC (v0.12.1). BAM files were generated using the GTEXv10 pipeline (https://github.com/broadinstitute/gtex-pipeline) using reference genome GRCh38 (Gencode, version 39). Briefly, FASTQ files were aligned using STAR (v2.7.10a) and optical duplicates marked by Picard Mark Duplicates (v3.3.0)[29]. Alignment metrics were assessed with RNA-SeQC (v2.4.2)[30]. Principal component analysis was performed using Transcript per million (TPM) values generated by RNA-SeQC and the prcomp function in R package stats (v4.4.0). Samples not clustering with other samples annotated to be derived from the same tissue source (either muscle, fibroblast or cycloheximide treated fibroblast) were removed (Fig S1).

### True Positive Definition

Diagnosed samples that passed quality control and contained diagnostic aberrant RNA events were manually reviewed in the Integrative Genomics Viewer (IGV)[31,32]. Each sample contributed one true positive gene for evaluation. Each true positive gene could be included as a true positive across more than one analysis type (*e.g*. splicing and expression) based on underlying mechanism. Splicing true positives were defined as containing ≥3 reads supporting a novel splice junction created by the pathogenic variant. Expression true positives were defined by a confirmed loss-of-function mechanism with normalized transcript abundance ≤50% or ≥ 100% of the corresponding normalized GTEx TPM. Genes were normalized to the geometric mean of *HPRT1*, *PPIA*, *YWHAZ* and *TBP*. Allelic imbalance true positives were defined by monoallelic loss-of-function events supported by single nucleotide polymorphism (SNP) analysis demonstrating a marked allelic ratio ≥70:30 at the site of the rare variant.

### True Negative Definition

Patients harboring pathogenic variants with no direct effect on splicing or expression (missense, in frame indels, terminal gene frameshifts) were considered as “true negatives” for RNA anomaly. RNA-seq data for these patients were manually reviewed in IGV to confirm that splicing was canonical and that no expression changes or allelic imbalance were present.

### Computational Tools

All tools were run on Biowulf (https://hpc.nih.gov). Current RNA-seq tools for rare disease use two main strategies: outlier-based (one-vs-all) and event-based (case-vs-control) approaches [33]. Outlier-based methods assume that each rare disease is unique within a heterogeneous cohort, an assumption that holds for some muscle disorder cohorts. However, it was not a valid assumption for our dataset, which includes recurrent variants and genes. Additionally, small-batch sequencing within the cohort likely introduced variability across samples. Event-based tools that compare cases to matched controls were considered as a strategy to mitigate these issues. We attempted to evaluate both case-versus-control and one-versus-all strategies when possible; however, this was only possible for splicing analysis in practice.

### One vs All Splice Tools (Outlier based approaches)

Three popular tools—FRASER, FRASER2.0, and LeafcutterMD [20,23,34] —were selected for evaluation. Since these tools require ≥30 samples, we created sample groups by tissue of origin, mixing diagnosed and undiagnosed RNA-seq samples from the same tissue: muscle (n=71), fibroblasts (n=37), and cycloheximide-treated fibroblasts (CHX, n=79). Muscle samples were sequenced in multiple small batches over 14 years using different library preparations (Table S1), primarily from the quadriceps (Fig S2). Although FRASER recommends analyzing stranded and unstranded libraries separately, preliminary analysis showed higher true positive recall when both library preparation methods were combined (Fig S3E-F). Therefore, all three tools were run with mixed library preparation.

FRASER was executed via the DROP pipeline (v1.2.2), using default parameters and a false discovery rate (FDR) of 5% (Fig S3A-C) [24]. FRASER2.0 was similarly run via the DROP pipeline (v1.4.0) with standard settings and an FDR of 5% (Fig S3D) [24]. LeafcutterMD (v0.2.9) was applied with preprocessing of BAM files to generate intron clusters using Leafcutter, with recommended parameters (minimum 50 splice junctions per cluster and a maximum intron distance of 500 kb)[34,35]. Outlier detection was restricted to groups with at least 20 reads and a minimum intron cluster size of 50. Resulting junctions were annotated with Ensembl gene IDs using Bedops (v2.4.41), followed by conversion to gene symbols utilizing the annotationDBI package in R. Correction for multiple hypothesis testing was performed using a FDR of 5%.

### Case vs Control Splice Tools (Events-based approaches)

Two commonly used tools, rMATS-Turbo and MINTIE, were selected[19,22]. Each patient sample was run against a set of ten healthy control samples. We used ten samples for the control cohort based on MINTIE recommendations. Control samples were selected from the GTEx database to match the tissue source when available (Table S2). Muscle control samples were filtered for RNA Integrity Number (RIN) > 8, ischemic time < 1000 minutes, and absence of autolysis. Fibroblast controls were selected based on tissue type "skin" and tissue type detail "cells – cultured fibroblasts," further filtered for RIN = 10 and ischemic time < 400 minutes. Due to the lack of available disease-free controls for CHX samples, case vs controls approaches were not tested.

MINTIE (v0.4.1) was run using FASTQ files from each patient sample against the ten selected GTEx controls, with recommended parameters and additional settings (splice_motif_mismatch = 3, FDR = 5%)[22]. Output was filtered to include only splice-related outliers, excluding large deletions, insertions, and other structural variants. rMATS-Turbo (v4.1.2) was executed on BAM files, with each patient sample compared to ten controls[19]. Standard parameters were used, with additional flags --novelSS and --allow-clipping. The maximum exon length for novel splice isoforms was set to 700 bp (-mel 700) to accommodate detection of large pseudoexons based on the largest in our true positive set. Correction for multiple hypothesis testing was performed using an FDR of 5%. All five MATS output files were included for assessment of true positive events, regardless of whether the detected aberrant splice category matched the expected event (e.g., intronic cryptic donor activation detected as intron retention).

### Expression Tools

OUTRIDER was run on all three cohorts as previously described in a one vs all manner via the DROP pipeline (v1.2.2) with recommended parameters which included an FDR of 1%[21,23]. Since OUTRIDER is designed as a one vs all approach, we performed a second run for the muscle patient samples with an addition of 100 control muscle samples downloaded from GTEx to test if the influx of healthy samples improves detection (Table S2).

### Allelic Imbalance Tools

The approach to detecting allelic imbalance in transcriptome data relies on heterozygous presence of variants in DNA. Thus, we restricted our test set to muscle samples where exome or genome variant call files (VCFs) were readily available, for a subset cohort of 59 samples. Two tools were selected, MAE module in the DROP pipeline [24] which uses variant level allele specific expression (ASE) and ANEVA-h, which uses gene level ASE[36]. The MAE module in DROP (v1.4.0) was run using standard parameters except modification of the allelic balance threshold to 70 instead of 80, reflecting our true positive dataset[9,24]. ANEVA-h required preprocessing of VCF and BAM files as follows. Haplotype-resolved, gene-level ASE data were generated for each case to enable regulatory outlier detection using ANEVA-h. RNA-seq libraries were processed and aligned to the GRCh38 reference genome as previously described. Corresponding genotype data were obtained from matched genome sequencing (GS) and phased using EAGLE (v2.4.1), leveraging the 1000 Genomes Project Phase 3 (30x) high-coverage reference panel for population-based phasing[37,38]. Phased genotypes and aligned RNA-seq BAM files were used as input to phASER (v1.1.1), which was run following best practices for haplotype-resolved ASE quantification [39]. Gene-level ASE values were computed using the phaser_gene_ae.py module, with heterozygous exonic SNPs aggregated by haplotype using GENCODE v39 transcript annotations [39]. ANEVA-h was then applied using the ANEVADOT_test function from the ANEVA-DOT R package, with default parameters [40]. Analyses were performed using reference VG estimates derived from 706 skeletal muscle RNA-seq samples from GTEx v8, ensuring tissue-specific calibration ( https://zenodo.org/records/15226130, https://zenodo.org/records/15226575). A minimum coverage threshold of 10 reads per gene was applied for inclusion. Multiple testing correction was performed using the Benjamini–Hochberg FDR procedure, and genes with FDR-adjusted p-values < 0.05 were designated as significant ASE outliers.

### True Positive and Negative Detection

Tool performance was evaluated based on detection of known aberrant RNA events. For splicing, a correctly detected true positive was defined by an exact coordinate match to the validated splice event. For expression, the true positive gene had to appear among reported outliers. For allelic imbalance, MAE required inclusion of the correct rare variant, while ANEVA-H required inclusion of the corresponding gene. True negatives were assessed by the absence of reported outliers, with correct detection defined as no outliers identified in the sample.

### Performance Metrics

Confusion matrices for True Positive (TP), True Negatives (TN), False Positives (FP) and False Negatives (FN) were established for each tool, allowing calculation of Precision, Accuracy, Recall, False Positive Rate (FPR), F1 score, and Mathews Correlation Coefficient (MCC).

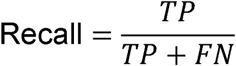

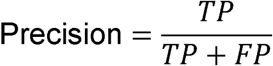

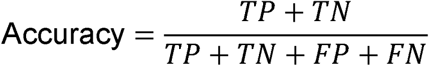

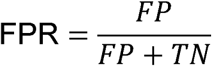

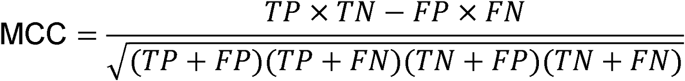

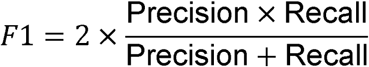

### Variant Annotation

Individual VCFs generated from exome sequencing and genome sequencing were annotated for population allele frequencies and splice prediction scores using the VEP_annotate tool (v113)[41]. SpliceAI scores were obtained as precomputed datasets from Biowulf[42]. Allele frequencies for each variant were sourced from gnomAD genomes v4.0 (https://gnomad.broadinstitute.org/).

Subsequent filtering of annotated VCFs was performed using VEP_filter (v113) to retain only variants with an allele frequency less than 0.01 or 0.0005 (a high and low threshold relevant to rare disease to assess how different cutoffs affect final outlier number) and with no homozygotes reported in gnomAD. Filtered VCFs were then converted to BED format using Bedops (version 2.4.41 megarow) with the convert2bed function.

### Outlier Evaluation

For splicing, an ensemble approach with variant filtration was used. Chromosomal coordinates for each outlier splice event were extracted into a master bed file (see Supplemental Methods). The master bed file was sorted using bedops (v2.4.41) and intersected with the filtered annotated variant bed file using the --range -50:50 -e set operation to identify variants overlapping splice outliers. Variants that intersected the outlier bed file were then filtered for spliceAI score ≥ 0.1, segregated in seqr and manually evaluated with the integrative genomics viewer (IGV) [32,43] (see Supplemental Methods). For OUTRIDER and ANEVA-h, the number of variants identified per individual was sufficiently low to allow for manual inspection of all events without additional filtration.

Candidate gene variants identified for undiagnosed patients were submitted to GeneMatcher[44].

## Results

### Truth set selection

We used RNA-seq data from 97 previously diagnosed patient samples to benchmark RNA-seq analysis tools. To ensure RNA-seq samples were correct, each BAM file was confirmed to harbor the known pathogenic variant in IGV. RNA-seq samples with independently verified variants affecting RNA were manually validated to contain evidence of aberrant splicing, expression or allelic imbalance in IGV. This set was defined as our true positive set (76 samples from 68 probands, Fig 1A,C, see methods for detailed definition). RNA-seq samples with confirmed pathogenic missense variants, inframe indels and premature truncating variants without nonsense mediated decay (NMD) and not expected to cause RNA changes were manually confirmed to contain no visible transcriptome changes in the pathogenic gene. This set was defined as our true negative set (21 samples, Fig 1A, see methods for detailed definition).

**Figure 1:**
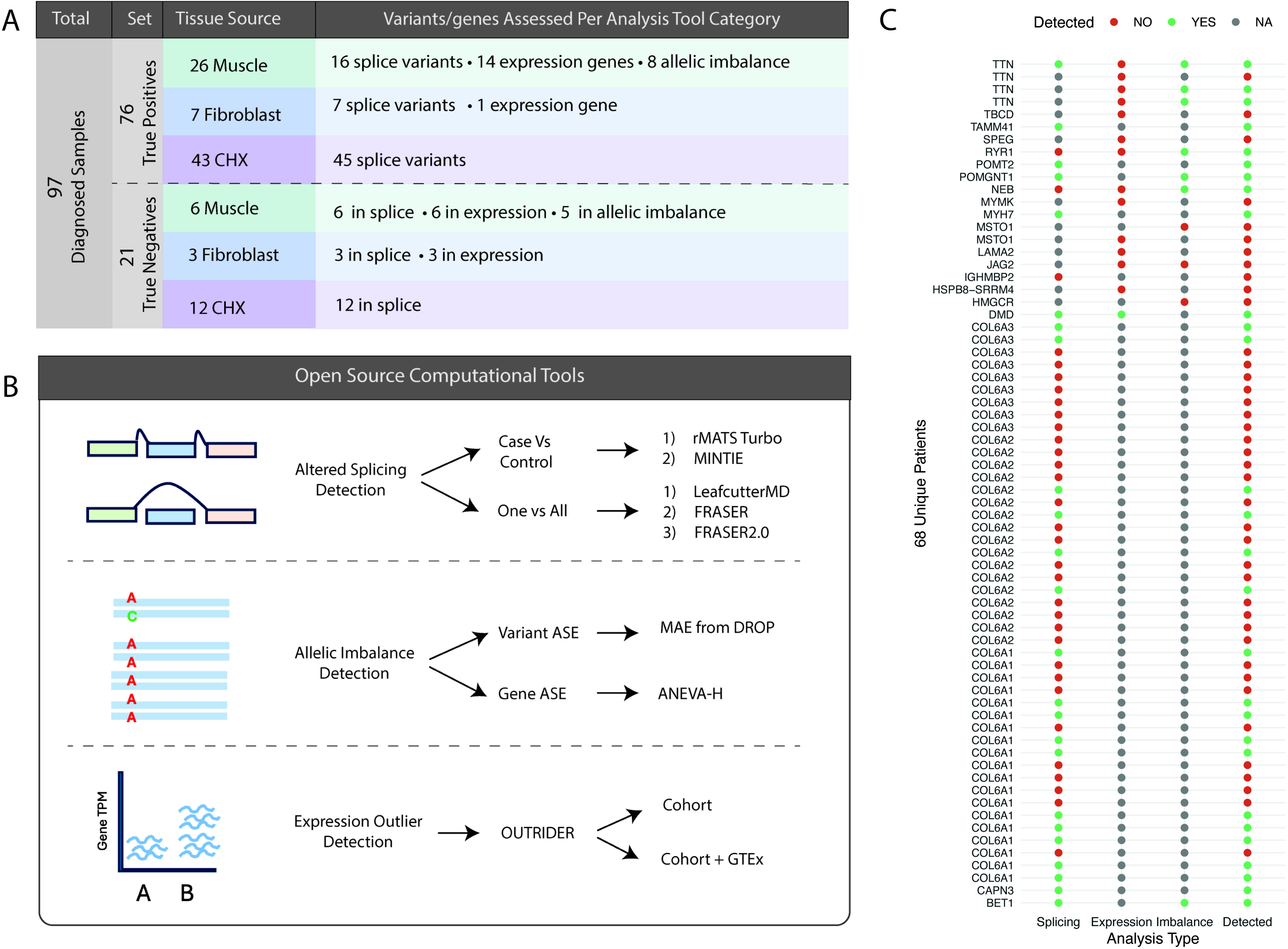
Truth cohorts included for analysis, workflow design and overall results. A) Truth set comprises of 97 samples, split into true positive and true negative sets. True positives and negatives were further categorized by tissue of origin and by the number of variants or genes contributed to each analysis type B) Benchmarking was performed on the eight tools with diverse methodolgies covering three RNA analysis approaches: aberrant splicing, allelic imbalance and expression outlier detection (ASE = allele specific expression). C) A total of 76 true positive samples from 68 probands with pathogenic variants leading to confirmed RNA events. Probands have causative variants leading to more than one aberrant RNA event type. Each proband is reported here by genetic diagnosis along with which analysis type the event was considered a true positive for. Red dots for undetected event, green dots for correct detection and grey dots when the sample was not tested.

### Recall rate of splice tools identifies ensemble strategy as most sensitive

To determine the performance of splice tools, we compared five tools using both case–control and one-vs-all designs to recall true positive splice events by tissue source (68 true positive variants from muscle, fibroblast and cycloheximide treated fibroblasts, Fig. 1A-B). In muscle derived true positive samples, case–control tools were most sensitive: rMATS (56%) > MINTIE (37.5%) > FRASER (33%) > FRASER2.0 (12.5%) > LeafcutterMD (0%) (16 true positive variants, Table S3, Fig. 2A). FRASER2.0 was excluded from further analysis as it underperformed FRASER. In fibroblast derived true positives, rMATS again performed best (86%) > MINTIE (28%) > FRASER (14%) > LeafcutterMD (0%) (7 true positive variants, Table S4, Fig 2A). Cycloheximide (CHX) treatment of fibroblasts can halt NMD by blocking protein synthesis, increasing detection of degraded aberrant events. In CHX-treated fibroblast true positives, only outlier-based methods were tested due to lack of control data, FRASER detected 33% and LeafcutterMD none (45 true positive variants, Table S5, Fig 2A).

**Figure 2:**
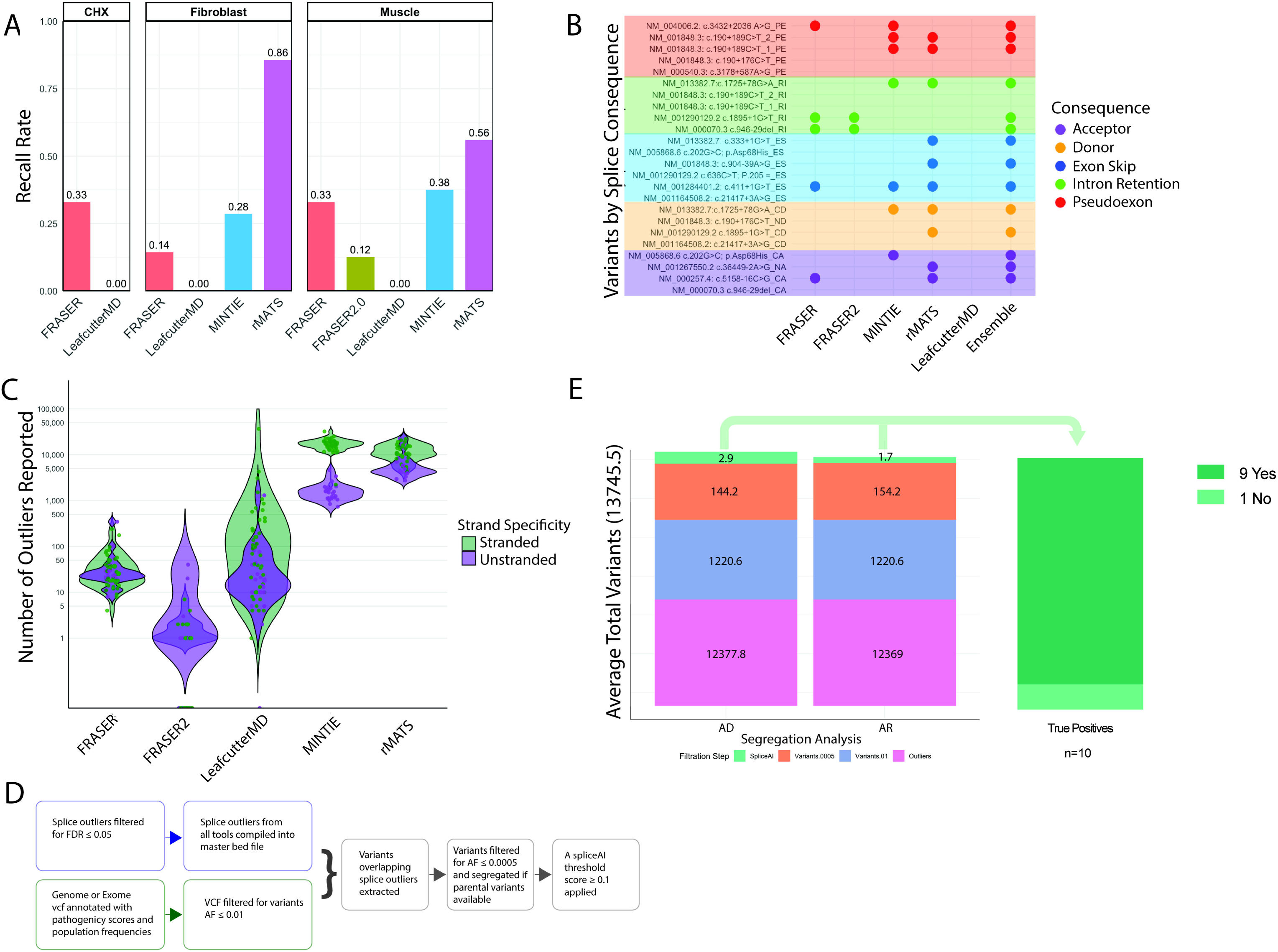
Results of true positive aberrant splice analysis. A) Recall rates of true positive splice variants for rMATS, MINTIE, LeafcutterMD, FRASER and FRASER2.0, broken down by tissue of origin. FRASER2.0 was only tested in the muscle set. For CHX set, only LeafcutterMD and FRASER were evaluated. Muscle = muscle-derived RNA-seq (16 true positive variants), Fibroblast = fibroblast derived RNA-seq (7 true positive variants), and CHX = cycloheximide-treated fibroblast derived RNA-seq (45 true positive variants) B) True positive splice variants create more than one aberrant splice event. Splice events were categorized as follows: donor event (CD = cryptic donor, ND = novel donor), acceptor event (CA = cryptic acceptor, NA= novel acceptor), exon skip (ES), pseudoexon (PE) and retained intron (RI). To evaluate any splice category preference among tools, each splice event from muscle true positive set is reported clustered by category with a dot if correctly detected by corresponding tool. An ensemble approach detects the most events. C) Violin plots showing the number of outliers reported for each sample by each tool in the muscle set, colored by library preparation differences. D) Variant based filtration strategy: outliers from all tools were merged into an ensemble file and intersected with rare DNA variants to reduce outlier burden, reporting outliers associated with rare variants only. E) Out of 16 true positive splice variants in the muscle set, 11 variants correctly detected (arising from 10 probands) were evaluated with variant filtration. Outliers were aggregated across all splice tools, excluding LeafcutterMD (average 13745.5 outliers/sample). Two allele frequencies were evaluated to assess maximum and minimum final outliers after variant based filtration. A lenient threshold representing the maximum variant allele frequency used for rare disease analysis (AF<0.01) with average outlier-associated variants after filtration 1220.6. Variants were then segregated, searched for autosomal dominant (AD) or autosomal recessive (AR) inheritance patterns and restricted to a stringent allele frequency threshold commonly applied in rare disease (AF<0.0005), reducing outliers to an average of 144.2 and 154.2, respectively. A final filtration by SpliceAI score < 0.1 reduced outliers to 2.9 and 1.7, respectively. Across 10 probands, 9 cases were correctly included in the final outlier list.

To evaluate if underperforming tools contributed unique detections, we examined recall by splice event category. FRASER detected two intron retention events, contributing one variant missed by other tools, supporting its inclusion (Fig 2B). MINTIE uniquely captured multiple splice events of a *BET1* variant (NM_005868.6: c.202G>C; p.Asp68His) in fibroblast and muscle tissue (Fig 2B, Tables S3-4). Although case-control methods have the highest recall rates, an ensemble approach aggregating outliers across multiple tools is most sensitive. In muscle, when tool output is combined, 68% (11/16) true positives are recalled, indicating greater overall yield with ensemble strategies (Fig 2B).

To understand the limitations of tool use, we examined the true positives undetected by any tool (Tables S3-5). These reflected common limitations: low mapping quality (*POMGNT1* NM_001290129.2 c.636C>T; p.205 =), few spanning splice junction reads due to degradation (*RYR1* NM_000540.3: c.3178+587A>G; *NEB* NM_001164508.2: c.21417+3A>G), complex loci with age specific alternative exon usage or highly repetitive regions (*NEB* NM_001164508.2: c.21417+3A>G, *IGHMBP2* NM_002180.3: c.1235+450 G>A), or inaccurate splice junction splitting (*RYR1* NM_000540.3: c.3178+587A>G *, COL6A1* NM_001848.3: c.190+176C>T, *IGHMBP2* NM_002180.3: c.1235+450 G>A). These cases demonstrate the continuing importance of manual investigation guided by clinical and genetic hypothesis.

Clinical application of splice detection tools requires a manageable number of outliers for review. We assessed output interpretability by comparing outlier counts reported on average per sample. In muscle, MINTIE and rMATS reported thousands of outliers, particularly when strand-specific data were compared to unstranded controls (Fig. 2C). In fibroblast and cycloheximide-treated fibroblast samples, outlier counts clustered by sequencing center, reflecting batch and library preparation effects (Figs. S4A–D).

To reduce outlier burden, we filtered for events overlapping rare DNA variants (Fig. 2D). Among ten true positive samples with genomic data, filtering for AF ≤ 0.01, a lenient cutoff for rare disease, yielded a mean of 1,221 variants per sample; applying a stricter threshold (AF ≤ 0.0005, segregation when available, and SpliceAI ≥ 0.1) reduced this to 1–3 variants (Figs. 2D–E, Table S6, Supplemental Methods). In nine of ten cases, the causal variant remained post-filtering. The missed case (*POMT2* NM_013382.7) the first variant (c.333+1G>T) associated splice event was not included in the master bed file (see Supplemental Methods), while a second intronic variant (c.1725+78G>A) was not captured by exome sequencing, highlighting the added value of including RNA-based variant calling (Table S6). Overall, rare variant filtration is successful at overcoming the challenge presented by the high number of outlier calls.

### Recall rate of expression tools is low

To determine the utility of expression analysis, we benchmarked OUTRIDER (15 true positive genes, Fig 1A-B). Across muscle true positive cases the recall rate was 7%, and the addition of 100 healthy control samples did not improve the recall (14 true positive genes,Table 1, Table S7). Fibroblast true positives were not detected at all, and no true positives were assessed from the CHX samples (1 true positive gene, Table 1, Table S7). Overall, the combined recall rate was 6.6% (1/15), and inclusion of expression analysis did not identify diagnoses missed by other analysis types (Fig 1C). The average number of outliers reported per sample were low (Table 1). Notably, the inclusion of control samples increased the average number of outliers while not providing additional diagnostic benefit (Table 1).

**Table 1:** Results of true positive expression analysis across Muscle, Fibroblast and Cycloheximide (CHX)-Treated Fibroblasts sets. Muscle samples were run with and without an additional 100 healthy samples from GTEx.

| Cohort | Muscle | Muscle + GTEx | Fibroblast | CHX |
| --- | --- | --- | --- | --- |
| Sample size | 71 | 171 | 37 | 79 |
| Recall | 1/14 (7%) | 1/14 (7%) | 0/1 (0%) | NA |
| Avg Outliers/sample | 3.08 | 4.02 | 0.8 | 1.21 |

To understand the limitations of OUTRIDER, we evaluated the undetected true positives. *TTN* underexpression recurs four times in our group of true positives and may not be considered statistically significant when using the one vs all approach. No patterns emerged for why any of the other true positives were missed (Table S7).

### Recall rate of allelic imbalance highlights ANEVA-H

To benchmark allelic imbalance detection, we tested the MAE module from DROP and ANEVA-H (8 true positive genes, Fig. 1A-B). Both detected 5 of 8 true positives (62.5% recall; Fig. 3A, Table S8). The tools overlapped on four variants, each identifying one unique event, for a combined recall of 75% (6/8). Allelic imbalance analysis yielded 4 diagnoses missed by splice or expression outlier methods (68 total diagnoses, Fig. 1C), highlighting its complementary diagnostic value. ANEVA-H reported markedly fewer outliers per sample than MAE (mean 44 vs 167; Fig. 3B), supporting its clinical adaptability.

**Figure 3:**
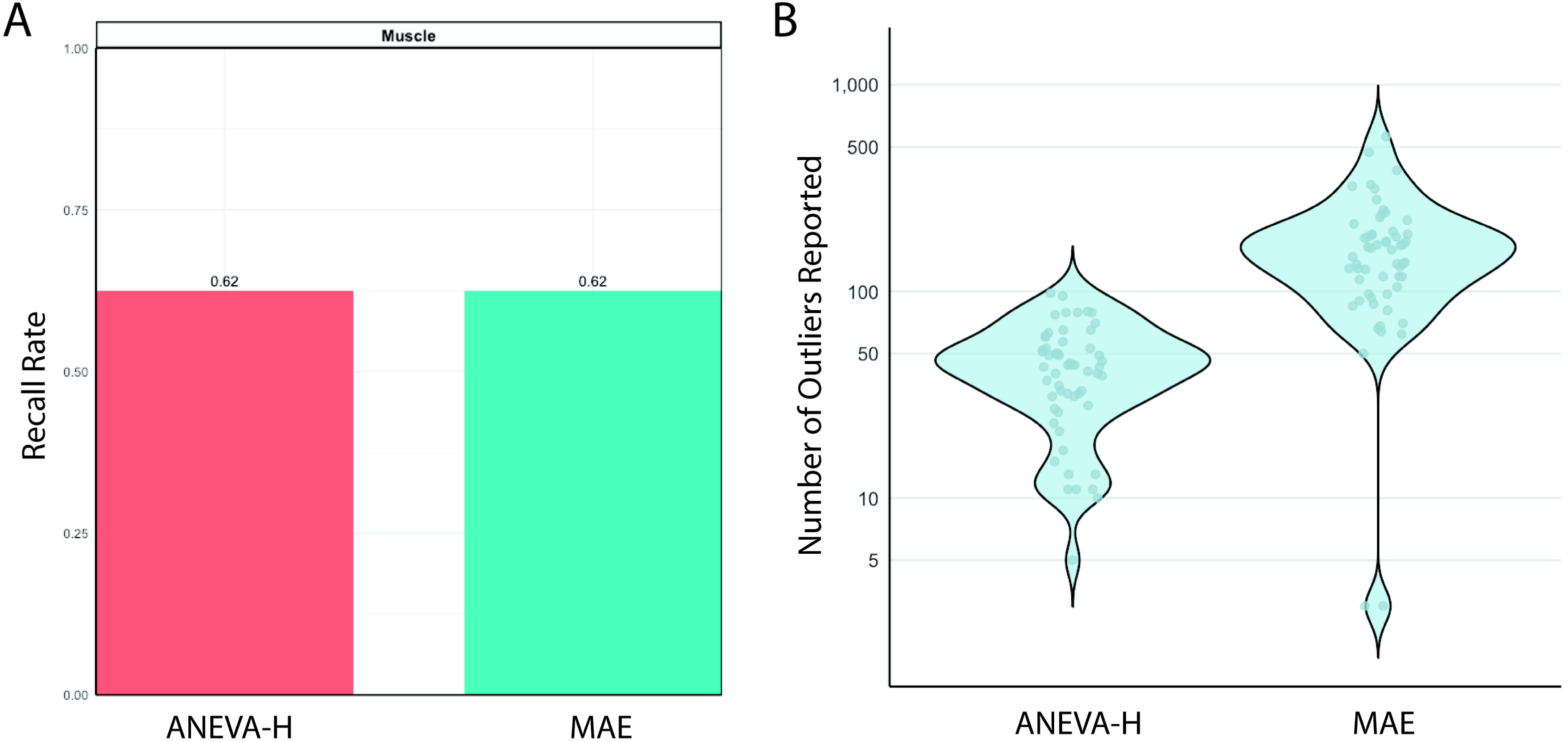
Results of true positive allelic imbalance analysis. A)Recall rates of 8 true positive genes for both tested tools, ANEVA-H and MAE. True positives were only tested in muscle derived RNA-seq samples B) Number of outliers reported for each sample per tool (59 total samples run).

Undetected cases involved *MSTO1*and *JAG2*. *MSTO1 (*NM_018116.4:c.1433A>G; p.Tyr478Cys & deletion) mapping is confounded by a pseudogene, and *JAG2 (*NM_002226.5: c.2515G>A; p.Gly839Arg & large deletion) was listed as homozygous in the VCF, masking monoallelic expression (Table S8). Two true positives were only detected by one tool each. ANEVA-H specifically missed *RYR1* (NM_000540.3: c.14126C>T; p.Thr4709Met & c.3178+587A>G) due to absent haplotype-specific SNPs in *RYR1* (required for gene level ASE generation), and MAE specifically missed *TTN* (NM_001267550.2: c.35794G>T; p.Glu11932Ter & c.22973C>A; p.Ser7658Ter) because of its size and repetitive regions (Table S8). Gene-level estimates from ANEVA-H may be more sensitive to subtle effects than variant-level MAE output.

### True negative analysis reveals high false positive rates

Unnecessary evaluation of RNA outliers in patients whose final diagnoses do not involve RNA-mediated mechanisms can divert limited clinical resources. To estimate how often such outliers occur, twenty-one true negative (defined in methods) RNA-seq samples were analyzed across all computational tools for presence or absence of outlier calls, as presence of any number of outliers will require clinical evaluation without benefit (Fig 1A). Among splicing tools, rMATS, MINTIE, LeafCutterMD, and FRASER reported outliers for all true negative samples (100%), while FRASER2.0 had a lower rate (33%) (Fig. 4A, Tables S3-5, Fig. S5). For expression tools, OUTRIDER with GTEx controls showed higher false positive rates than OUTRIDER alone (83% vs. 56%), confirming that OUTRIDER alone is both more sensitive and specific (Table S7, Fig 4A). For allelic imbalance tools, MAE produced more false positives (100%) than ANEVA-H (80%), further supporting ANEVA-H as the better-performing method given its lower outlier burden (Table S8, Fig. 4A).

**Figure 4:**
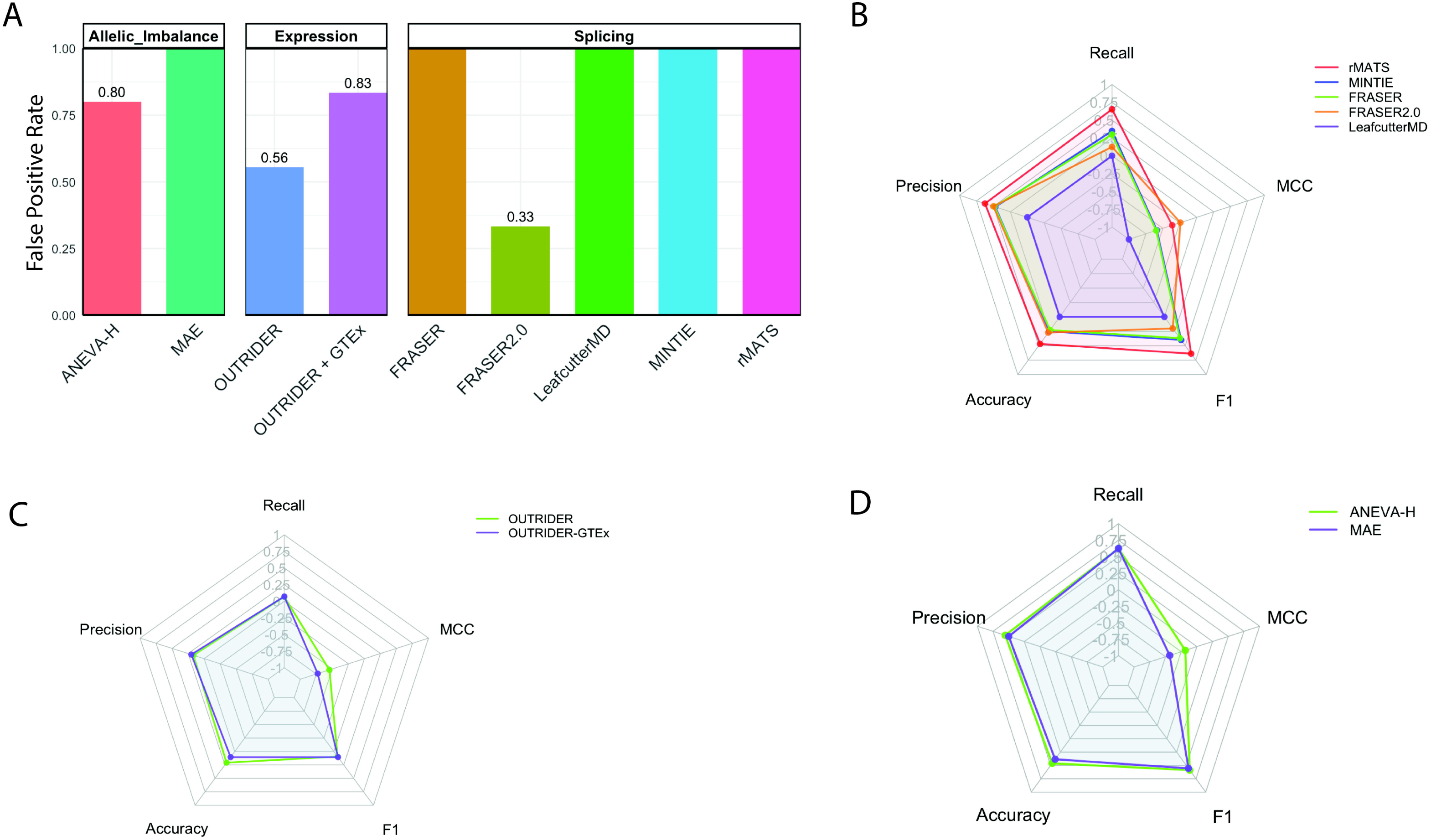
Precision, accuracy and predictive power. A) True negative samples (n=21) were evaluated across all tools by presence or absence of reported outliers. Results were aggregated across tissue sources to quanity the false positive rate for each tool tested, with 100% false positive rate reported when all true negative samples had outliers. Bar plots show the false positive rate for each tool, categorized by analysis subtypes splicing (rMATS = 9, MINTIE =9, FRASER = 21, FRASER2.0 = 6, LeafCutterMD=21), expression (OUTRIDER =9, OUTRIDER+GTEx =6) and allelic imbalance (ANEVA-H =5, MAE =5). B) True positve samples were similarly aggegated across tissue sources for each tool, allowing for determination of true positives, false positives, true negatives and false negatives for each tool (Table S9). Accuracy, precision, recall, F1 score and MCC were calculated. Radar plot show metrics for splice tools C) for allelic imbalance tools and D) for expression tools.

### Poor predictive performance across tools

To evaluate tool accuracy and precision, true positive and true negative detections were aggregated across tissue types for each tool (Table S9). Among splicing tools, rMATS achieved the highest precision, accuracy, recall, and F1 score (Fig. 4B, Table S9). For expression analysis, OUTRIDER alone outperformed the OUTRIDER + GTEx combination across all metrics (Fig. 4C, Table S9). In allelic imbalance analysis, ANEVA-H exceeded MAE on every metric (Fig. 4D, Table S9). Notably, Mathews Correlation Coefficient (MCC) values were negative across all tools, indicating limited overall predictive power (Fig 4B-D, Table S9). Across all analyses, only 28 of 68 (41%) known diagnoses were correctly detected (Fig. 1C), consistent with the MCC scores.

### Application to undiagnosed samples

We applied the top performing approaches integrating ensemble splice analysis with variant filtration, OUTRIDER, and ANEVA-H to 74 undiagnosed samples (Fig. 5A). In 28 muscle-derived samples, splice analysis identified 129 candidate variants (mean 4.6 per sample), of which 39 showed clear aberrant splicing on manual review. Nine variants were prioritized for further evaluation after segregation and analysis for a possible second hit if not de novo (Figs. 5B-C, Table S10). None were located in OMIM-listed disease genes. OUTRIDER analysis identified one candidate in a CHX treated sample, *TRIP4*, with reduced expression, in a patient with phenotypically compatible disease (Fig. S6A–B); however, exome, genome, and optical mapping revealed no causal variants, and follow-up RNA-seq confirmed normal expression and splicing in untreated fibroblasts (Fig. S6C–D). ANEVA-H did not identify additional candidates. Together, these analyses demonstrate that splicing-based approaches contributed novel candidate findings, whereas expression and allelic imbalance analyses did not improve diagnostic yield.

**Figure 5:**
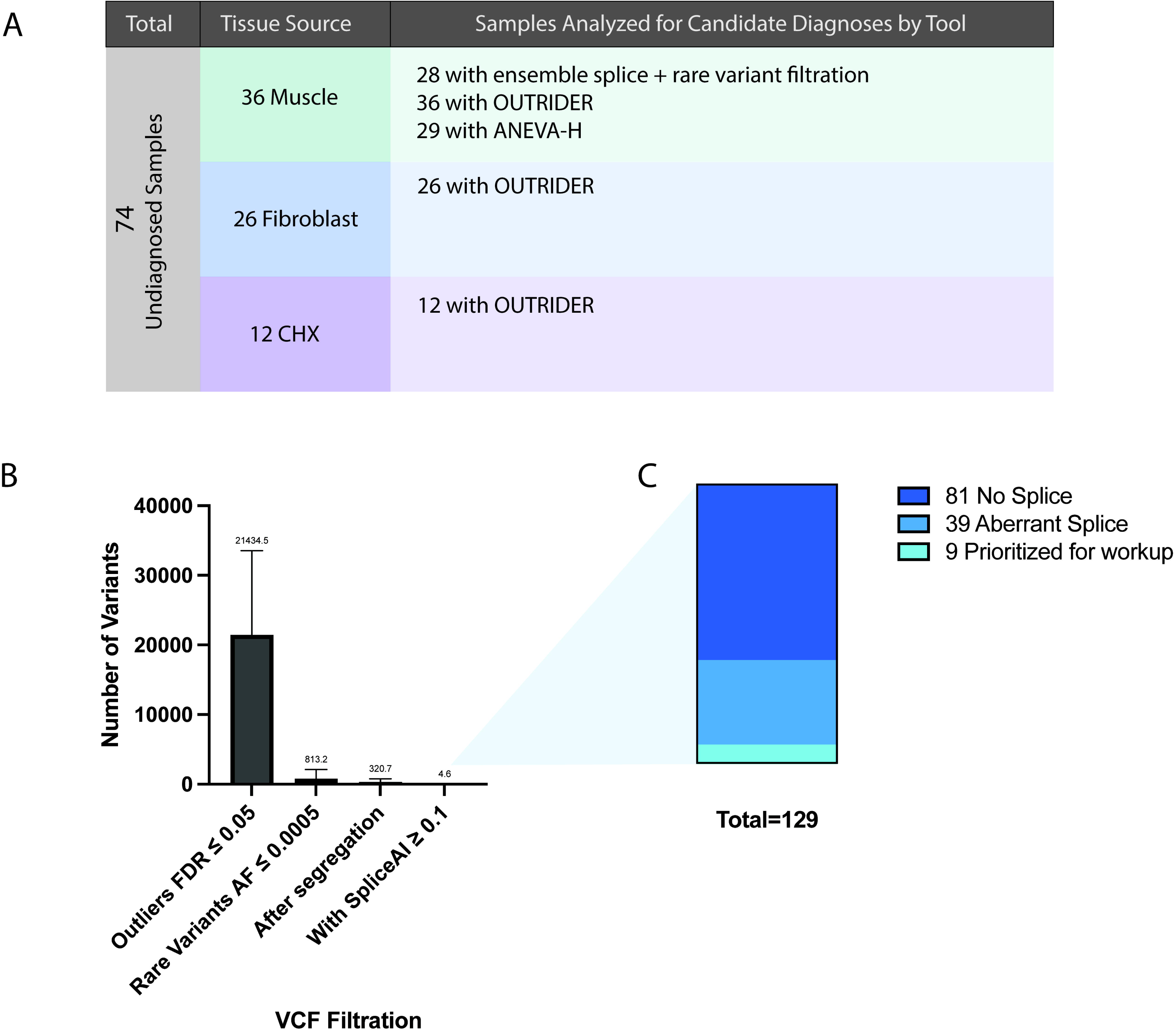
Application of RNA analysis to undiagnosed samples. A) 74 undiagnosed samples were evaluated by RNA analytical tools to identify candidate variants. Figure shows the numbers split by tissue source and list of analytical tools applied to each set of undiagnosed samples. B) For splice analysis, an ensemble approach was used along with filtration by rare DNA variants (AF < 0.0005). Bar plot shows subsequent steps of the filtration method in reducing the number of outliers to evaluate. C) 129 final outliers were manually evaluated across 28 patients, resulting in 9 final candidates submitted to geneMatcher.

## Discussion

Over a 14-year period, RNA-seq was integrated into our research diagnostic pipeline for patients with suspected Mendelian neuromuscular disorders, improving diagnostic yield but leaving a substantial fraction unresolved. To assess how transcriptome-centric methods could further enhance yield, we benchmarked commonly used RNA analysis tools.

Splicing tools consistently demonstrated the highest diagnostic sensitivity, detecting 24/68 diagnoses, in line with prior studies [5–7,14,15]. While outlier-based methods FRASER and FRASER2 are the most widely implemented, case–control approaches such as rMATS-turbo achieved greater sensitivity in our dataset. A prior report analyzing whole blood RNA-seq reported similar findings, while another report using muscle-derived RNA-seq found greater sensitivity with FRASER [5,7]. Differences in FDR thresholds applied to rMATS and FRASER may explain the discrepancy [5]. Although combining multiple tools improved recall in our dataset, it also increased outlier burden. Reported strategies to reduce outliers include restricting to clinically relevant genes or panels [5,6,15,16]. While this enhances interpretability, it does not consistently result in new diagnoses [15–17]. We chose not to restrict results this way given that our patients had undergone prior extensive phenotype based genetic analyses without success.

Treatment of cells with cycloheximide has been proposed to enhance detection of splice events subject to NMD, while the diagnostic yield reported has only been modest [15,17]. With only one sample recurring in our treated and untreated true positives, we could not perform comprehensive analysis to ascertain the diagnostic benefit of using CHX.

Expression outlier analysis assumes that loss-of-function (LoF) variants trigger NMD and reduce transcript abundance. Using OUTRIDER, with and without GTEx controls, we observed limited sensitivity and no novel diagnostic findings, consistent with previous reports [5–7,15,16]. Some outliers corresponded to known heterozygous pathogenic variants, while many others lacked supporting rare variants, likely reflecting pathway-level variation or common polymorphisms [6,14]. Tissue specificity and genome build inconsistencies may further confound interpretation [45,46]. Nevertheless, higher yields reported in other cohorts—such as NIH Undiagnosed Diseases Program fibroblast datasets[42,43] —suggest that expression analysis may perform better in phenotypically diverse or batch sequenced cohorts [47,48].

Allelic imbalance analysis remains underutilized in RNA-seq diagnostics, with only a few studies reporting its clinical application and modest diagnostic yield [5,14,16]. While variant level MAE and gene level ANEVA-H have similar sensitivity in our dataset, ANEVA-H produces fewer outliers and has a lower false positive rate. Genes reported as outliers by ANEVA-H frequently harbored structural variants or LoF variants, consistent with prior studies [49], yet lacked compatible inheritance or phenotypic correlation to make them candidate diagnoses. Imbalance analysis recovered four diagnoses in our true positive benchmarking missed by other tools, supporting its inclusion in RNA analysis pipelines. However, methods such as ANEVA-H remain constrained by haplotype phasing requirements, supporting ensemble approaches for now.

This study has several limitations. An optimal study would be prospective, using pre-defined criteria for negative and positive cases. However, our study required a minority subset (splicing and transcription modifying variants) of a set of rare diseases. We felt that a study of retrospective data, with pre-analytically defined case categorization, had a strong potential to generate useful information. That said, our findings would benefit from prospective implementation and replication. Additionally, variants with clinically interpretable mechanisms were selected for assessment of tools, reflective of downstream evaluation of outliers in the clinic. This favors clearly defined aberrant events and likely underestimates tool sensitivity, making discernment of a single tool’s superiority challenging. True negatives were evaluated by presence or absence of outliers to capture time spent assessing outliers for patients without underlying pathogenic aberrant RNA events. Evaluating true negatives by binary outlier classification may overestimate false positive rates, as some outliers may represent genuine biological variation rather than artifacts, and additionally, underestimates predictive ability of tools by lowering MCC scores.

Challenges in identifying clear diagnostic candidates in our 74 unresolved cases likely reflect both biological and technical factors. Firstly, unsolved cases may harbor non-RNA-affecting pathogenic mechanisms which would be missed by RNA analytical tools [42,50,51]. Secondly, muscle and fibroblast tissues may not be the relevant tissue for capturing pathogenic events in patients with neurogenic or neuromuscular junction phenotypes. Moreover, reliance on exome data in some cases excludes deep intronic variants, and addition of RNA variant calling or genome sequencing could provide more comprehensive detection.

Whether to apply RNA-centric versus RNA-complementary approaches remain a central question for diagnostic transcriptomics. Manual investigation of genomic variants in RNA-seq files is inherently complementary. In our benchmarking, RNA-based tools only recovered 28 of 68 known diagnoses with RNA events, and application of tools to undiagnosed cases required rare variant filtration, underscoring the interdependence of transcriptomic and genomic data with current technology. Similar trends have been reported across studies, where diagnostic gains stem largely from RNA evidence supporting previously identified VUSs or genes prioritized due to one pathogenic hit and phenotypic fit [5–7,15,16,27]. RNA-centric pipelines relying solely on RNA variant calls instead may miss pathogenic splice events when the variant is absent in the resulting transcript, while hybrid genome-plus-transcriptome strategies offer the most comprehensive diagnostic coverage [7,33,49,52]. Given the performance of the current tools, RNA approaches remain complementary to genomic analyses, with manual investigation still a necessary analytical tool.

## Conclusion

With currently available RNA-seq tools, we recommend using ensemble approaches in diagnostic pipelines rather than any single tool. Additionally, we recommend manual inspection of all VUS’s predicted to alter RNA even if no aberrant events are identified using tools. As RNA-seq continues to mature as a diagnostic tool, integrated genome-transcriptome pipelines, improved reference datasets, disease specific transcriptomic signatures and tissue-specific expression ranges will be essential for reducing false positives and enhancing interpretability [25,53]. Increased use of long-read RNA-seq will improve splice detection in difficult to map regions and genes with complex isoforms. Ultimately, such unified analytical frameworks will expedite variant prioritization and accelerate the path to molecular diagnosis.

## Data Availability

The datasets generated and/or analyzed during the current study are available in the dbGAP repository phs001272.v2.p1.

## List of Abbreviations

NMD: Nonsense Mediated Decay
CHX: Cycloheximide
ASE: Allele Specific Expression
IGV: Integrative Genomics Viewer
RNA-Seq: RNA-Sequencing
VUS: Variant of Uncertain Significance
LoF: Loss of Function
NIH: National Institutes of Health
SNP: Single Nucleotide Polymorphism
BAM: Binary Alignment File
AF: Allele Frequency
VCF: Variant Call File
MCC: Mathews Correlation Coefficient
TPM: Transcripts Per Million
FPR: False Positive Rate

## Declarations

### Ethics approval and consent to participate

Written informed consent and age-appropriate assent for study procedures were obtained by a qualified investigator [protocol 12-N-0095 approved by the National Institute of Neurological Disorders and Stroke, National Institutes of Health IRB].

### Consent for publication

All participating patients consented to publication of deidentified material

### Authors’ contributions

SS conceptualized the study design, performed all data analysis, and prepared the manuscript. KG ran the ANEVA-H data analysis and contributed manuscript preparation. SG contributed to the study design and manuscript preparation. SD performed initial genetic analysis for each patient used in the study and contributed to clinical interpretation of new candidate diagnoses. VB collected RNA from patients with COL6-RDs. JM ran FRASER and OUTRIDER on the cycloheximide treated RNA-seq samples. YH collected RNA from patient muscle and fibroblast for sequencing. PU contributed to study design and bioinformatic support. VG and BW contributed patient diagnoses used as true positives in the study. RO contribute to clinical evaluation of patients and interpretation of new candidate diagnoses. ARF contribute to clinical evaluation of patients. PM supervised ANEVA-H data analysis. DA supervised SS and conceptualized study design and manuscript preparation. CB supervised SS and conceptualized study design and manuscript preparation. All authors read and approved the final manuscript.

## Acknowledgements

This work utilized the computational resources of the NIH HPC Biowulf cluster (https://hpc.nih.gov).

With great thanks to referring physicians, patients and their families. We would like to thank Christopher Mendoza, Gilberto (“Mike”) Averion and Kia Brooks for their help and support in coordinating the patient evaluations.

## Supplemental Data

### Supplemental Tables

Table S1: Details on sequencing and library preparation across cohorts

Table S2: GTEx accession numbers for samples used in this study

Table S3: Muscle splicing true positives

Table S4: Fibroblast splicing true positives

Table S5: CHX-treated fibroblast splicing true positives

Table S6: Variant filtration of detected true positives in muscle

Table S7: Expression true positives

Table S8: Allelic imbalance true positives

Table S9: Confusion matrices and performance metrics

Table S10: Splicing candidate variants in 29 undiagnosed muscle samples

### Supplemental Figures

Figure S1: PCA analysis of cohorts

Figure S2: Breakdown of biopsy locations for muscle RNA-seq

Figure S3: FRASER parameters and preliminary tests

Figure S4: Outliers reported for splicing tools in fibroblast and cycloheximide treated fibroblast samples

Figure S5: Specificity rates per tool per tissue origin

Figure S6: Spurious find in cycloheximide treated fibroblast OUTRIDER analysis

