## Supplemental Figures S1-S6 for "Benchmarking RNA-seq Tools for Real-World Diagnostic Applications"

**Fig S1:** PCA analysis of cohorts

A

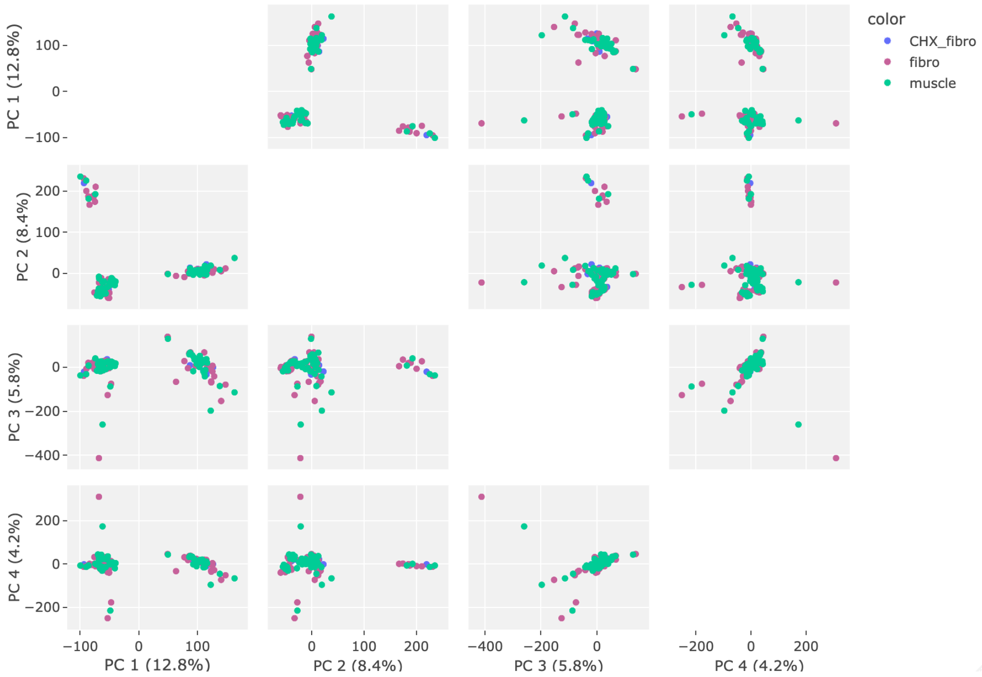


B

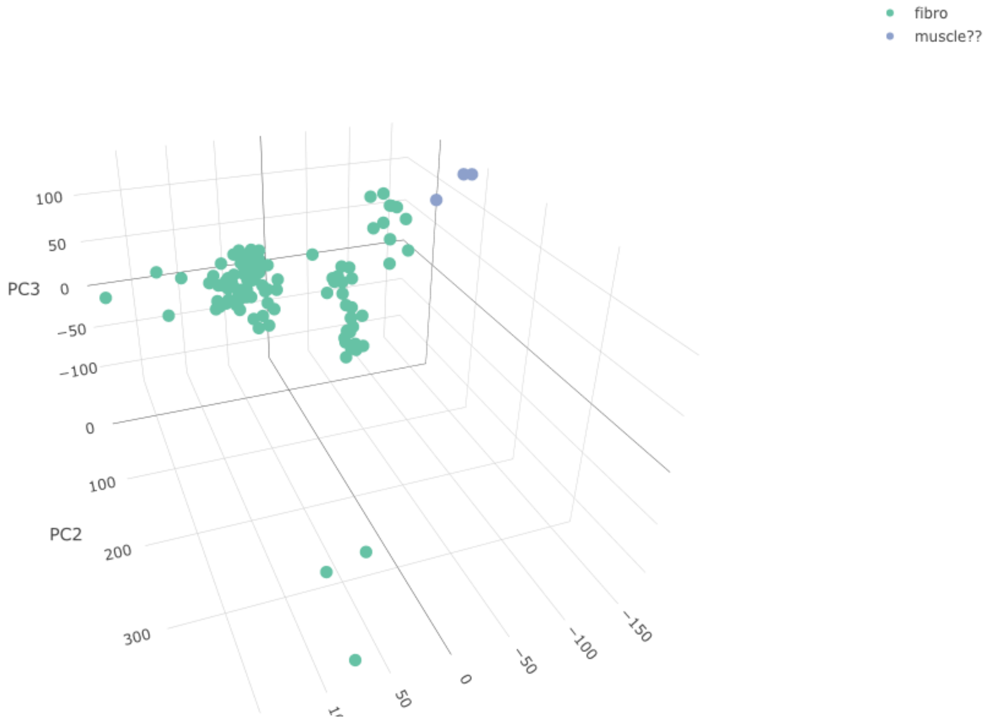


Figure S1: Principal component analysis (PCA) of RNA-seq samples annotated by tissue source. A) 2D PCA plot of top four principal components do not cluster by tissue source, rather by sequencing facility. B) 3D PCA plots of fibroblast samples show three samples annotated to be derived from muscle tissue clustering with fibroblast samples. These samples were removed.

**Fig S2:** Breakdown of biopsy locations for muscle RNA-seq

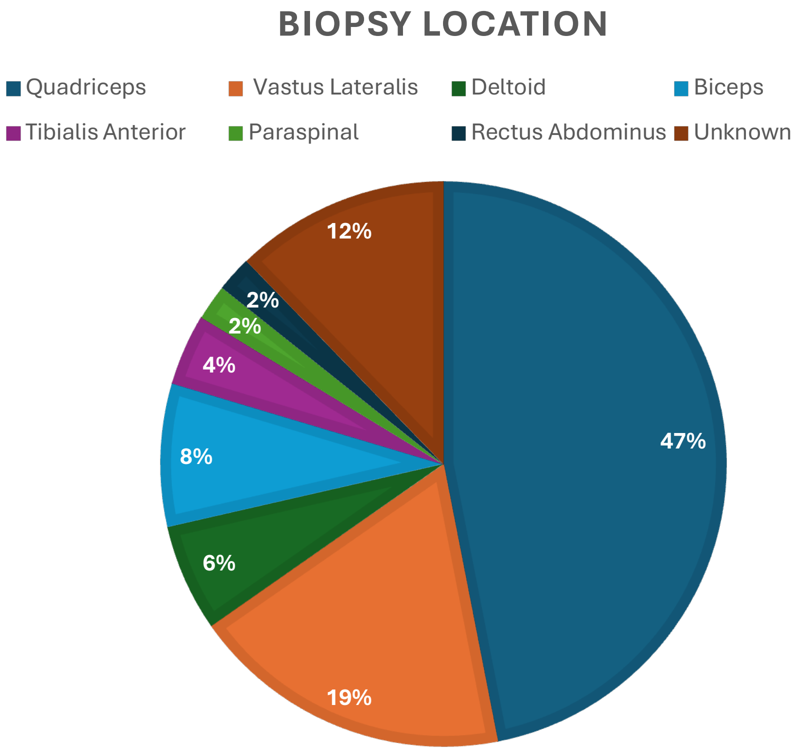


Figure S2: Muscle RNAseq samples split by biopsy location. Most of muscle samples were derived from quadriceps muscle (66%: 46% from quadriceps unspecified and 19% from vastus lateralis). The remainder were split between deltoid, biceps, tibialis anterior, paraspinals, rectus abdominus. 12% were unknown.

**Fig S3:** FRASER parameters and preliminary tests

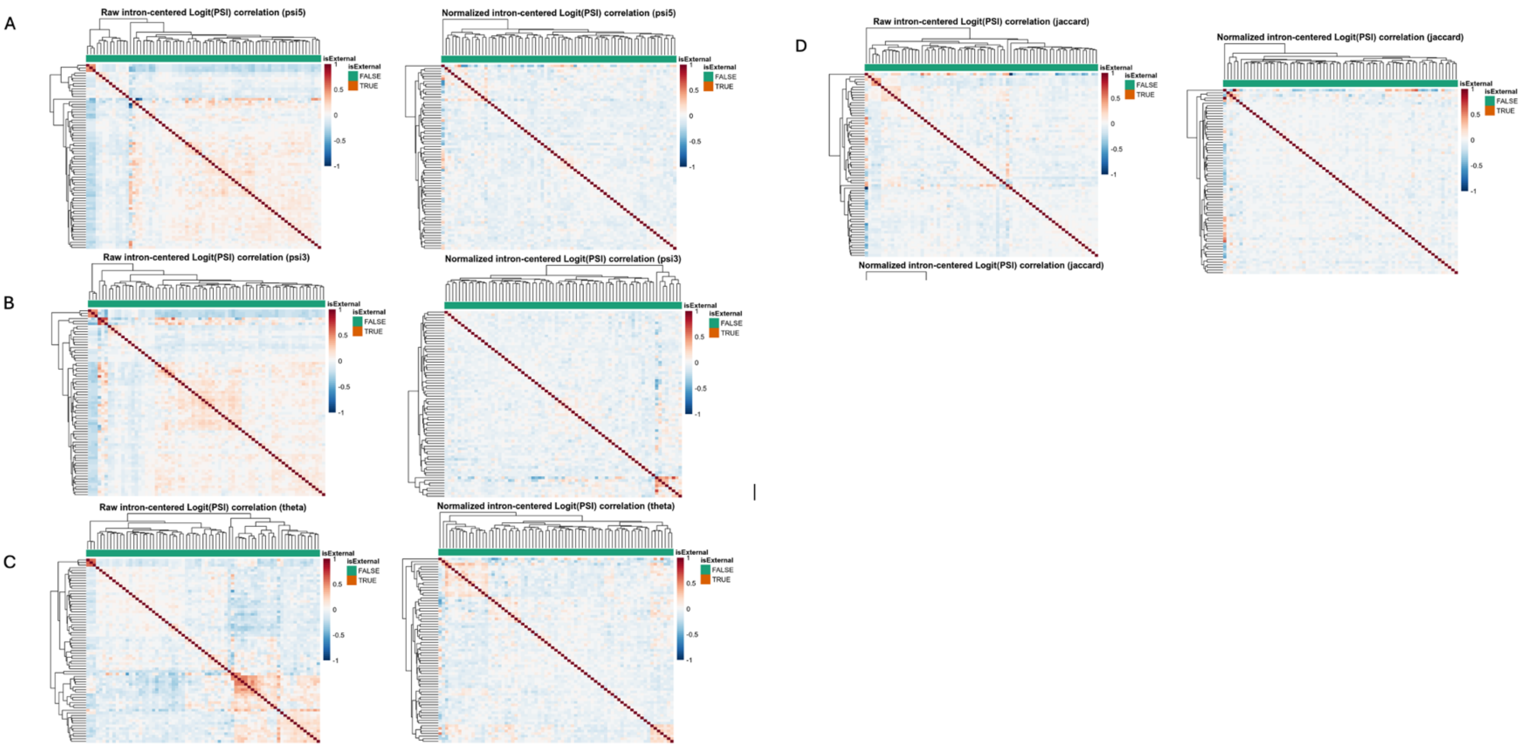


E

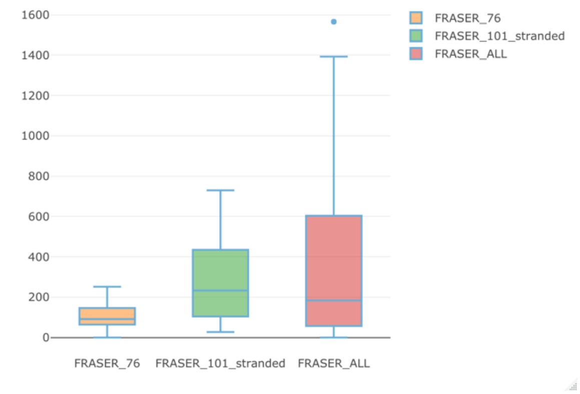


F

| Run | Total samples | True Positives | Correctly Detected |
|---|---|---|---|
| FRASER_76 unstranded | 20 | 6 | 1 |
| FRASER_101 stranded | 29 | 8 | 0 |
| FRASER_ALL | 49 | 14 | 2 |

Figure S3: Preliminary FRASER tests and denoising autoencoder. FRASER hierarchal clustering of samples pre and post (left and right) denoising autoencoder in A) Muscle samples B) Fibroblast samples C) Cycloheximide treated fibroblast samples. FRASER2.0 denoising pre and post in D) muscle. E) preliminary analysis of FRASER split by library prep methods and resulting number of outliers, with outlier number increased with increasingly diverse cohorts F) Breakdown of true positive recall by preliminary FRASER runs, although outliers are the highest with all samples combined, the highest recall rate is also noted.

**Fig S4:** Outliers reported for splicing tools in fibroblast and cycloheximide treated fibroblasts samples

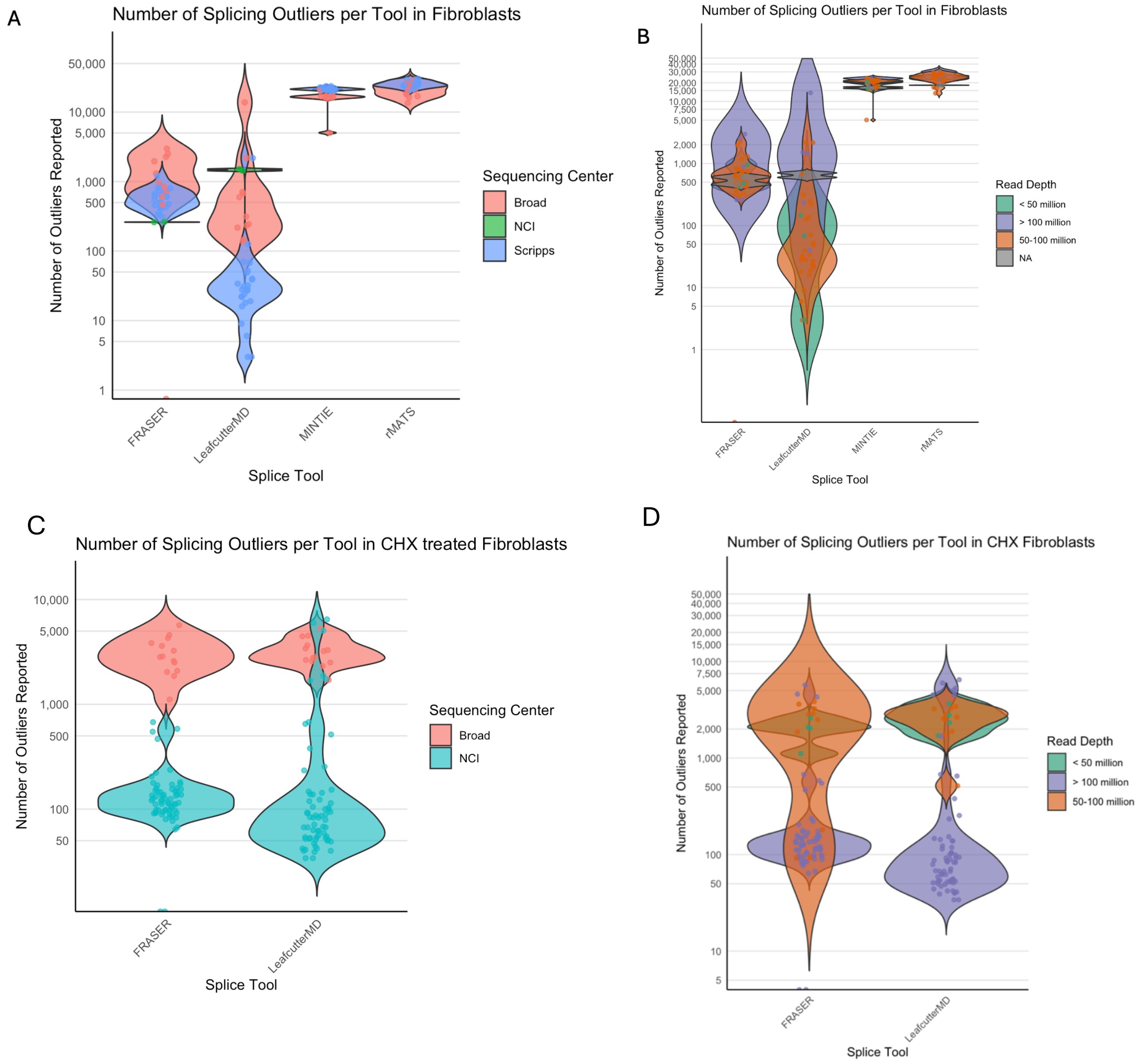


Figure S4: Outlier count violin plots for each tool split by tissue source origin. A) in fibroblast samples, the number of outliers cluster by sequencing center but not by B) read depth. In cycloheximide treated samples, outlier number again lusters by C) sequencing center but not by D) read depth.

**Fig S5:** Specificity rates per tool per tissue origin

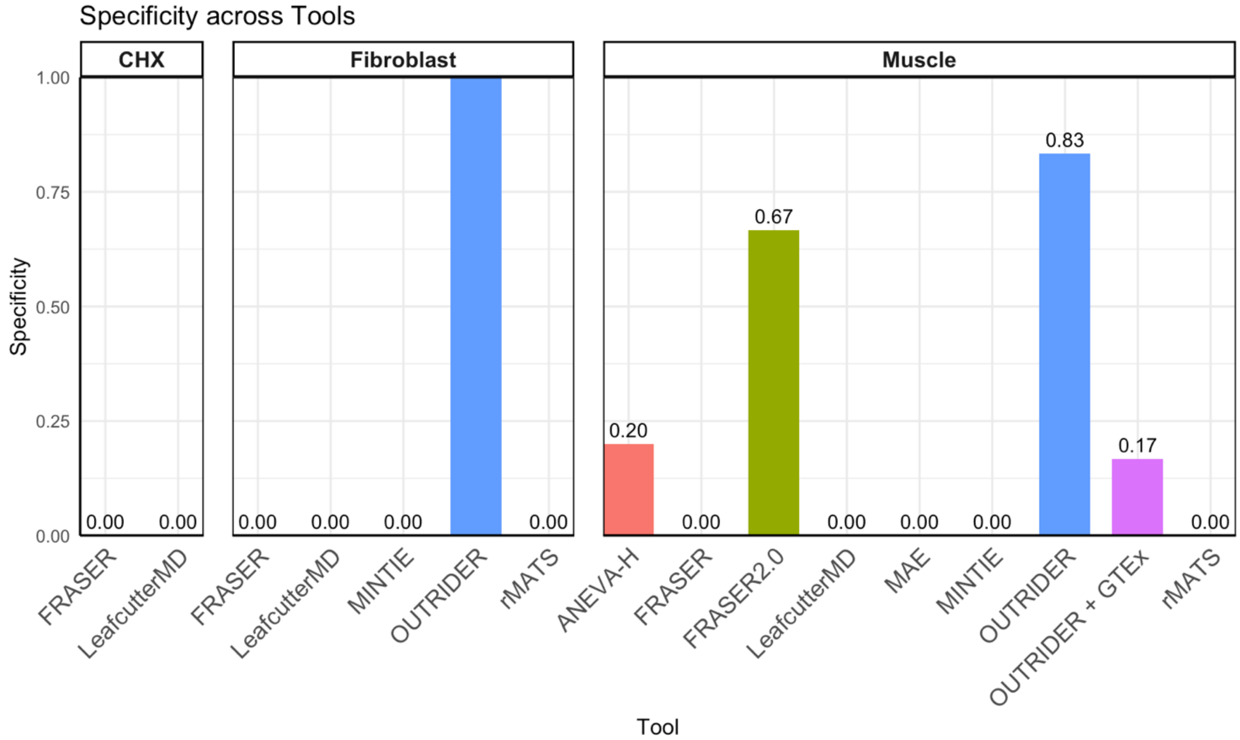


Figure S5: specificity rates for each tool per tissue. True negatives included in every tool were evaluated for presence or absence of outliers and reported here for every tool per tissue.

**Fig S6:** Spurious find in cycloheximide treated fibroblast OUTRIDER analysis

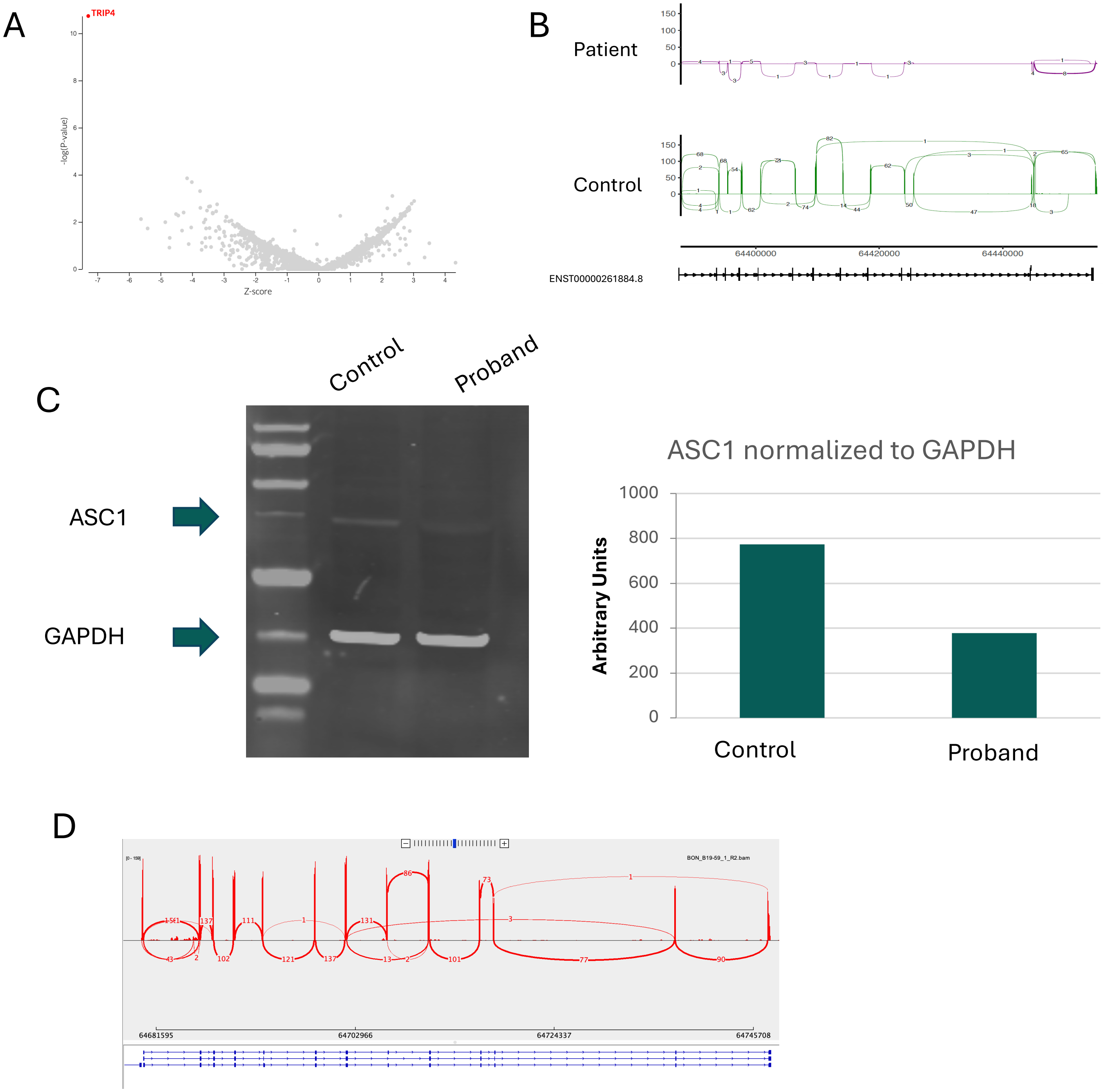


Figure S6: Workup of TRIP4 expression outlier from cycloheximide treated fibroblast cohort: A) TRIP4 identified as an expression outlier with OUTRIDER. B) manual inspection in IGV demonstrates almost no reads in patient over TRIP4 compared to controls (n=3, aggregate sashimi). C) ASC1 protein (encoded by TRIP4) is mildly reduced in patient compared to control in untreated fibroblasts D) Repeat RNAseq from untreated fibroblasts demonstrates normal TRIP4 expression.
