## Supplemental Methods for "Benchmarking RNA-seq Tools for Real-World Diagnostic Applications"

*DNA extraction*

DNA was extracted from blood, skin fibroblasts, saliva and the pelleted fraction of the urine specimen using the Gentra Puregene Blood kit (Qiagen, Germantown, MD).

*Exome Sequencing*:

Exome sequencing and data processing were performed by the Genomics Platform at the Broad Institute (Cambridge, MA).  Libraries from DNA samples (>250 ng of DNA, at >2 ng/ul) were created with an Illumina Nextera or Twist exome capture (~38 Mb target) and sequenced (150 bp paired reads) to cover >85% of targets at 20x and a mean target coverage of >55x. Sample identity quality assurance checks were performed on each sample. The exome sequencing data was de-multiplexed and each sample's sequence data were aggregated into a single Picard BAM file.

*Genome Sequencing:*

Genome sequencing and data processing were performed by the Genomics Platform at the Broad Institute of MIT and Harvard.  PCR-free preparation of sample DNA (350 ng input at >2 ng/ul) is accomplished using Illumina HiSeq X Ten v2 chemistry. Libraries are sequenced to a mean target coverage of >30x.

*Exome/Genome Alignment and Variant Calling*

Exome and Genome sequencing data was processed through a pipeline based on Picard, using base quality score recalibration and local realignment at known indels. The BWA aligner was used for mapping reads to the human genome build 38. Single Nucleotide Variants (SNVs) and insertions/deletions (indels) are jointly called across all samples using the Genome Analysis Toolkit (GATK) HaplotypeCaller package version 4.0. Default filters were applied to SNV and indel calls using the GATK Variant Quality Score Recalibration (VQSR) approach.

*RNA Extraction*

RNA was isolated from 10mg of frozen muscle biopsy using QIAzol (Qiagen, Germantown, Maryland, USA) manufacturer’s instructions. The aqueous phase was isolated after addition of chloroform and the RNA was dissolved in ethanol and cleaned on spin columns and eluted in RNase free water. RNA was isolated from patient derived skin fibroblast cultures using the Qiagen miRNeasy kit following the manufacturer’s protocol. RNA was evaluated using a BioAnalyzer tape station.

*Cycloheximide treatment*

For samples sequenced in the COL6-RD cohort, Fibroblasts were seeded at 2x10^5^ in 6 well plates and cultured in high glucose DMEM with 10% FBS and 1% penicillin-streptomycin. The following day, each well received 150ug/ml cycloheximide treatment (Sigma, Saint Louis, Missouri for 5 hours, and subsequently collected in trizol for RNA extraction.

For remaining samples in the cycloheximide treated cohort, patient dermal fibroblasts were treated with 200ug /ml cycloheximide) in 6-well plate for 19 hours and processed as described above.

*RNA sequencing*

Human bulk transcriptome sequencing was performed by the Genomics Platform at the Broad Institute of MIT and Harvard, NIH core facilities (NCI), and Scripps Research of La Jolla. The transcriptome product combines poly(A)-selection of mRNA transcripts with a strand-specific cDNA library preparation, with a mean insert size of 550bp. Some samples were prepared without strand specific protocols. Libraries were sequenced on the HiSeq 2500 platform to a minimum depth of 50 million reads. ERCC RNA controls are included for all samples, allowing additional control of variability between samples.

*Muscle sequencing QC*

Muscle samples were all sequenced at Broad, using a mixture of stranded and unstranded protocols as described above (Table S1). Samples were all on average 80M reads.

*Fibroblast sequencing QC*

The fibroblast derived RNAseq samples were sequenced at three different facilities. All samples were prepared using polyA selection and strand-specific protocols; however, read depth and read lengths varied (Table S1). Two samples sequenced at the NCI facility had an average read depth of 175M, samples from Broad had an average read depth of 80M reads, while samples from Scripps averaged 60M reads per sample. The number of genes detected was similar across the second two cohorts, ranging from 19,000 to 21,000 (Supplemental Figure 4B). The number of genes detected was higher from the NCI sequencing center, ranging from 22,000-24,000.

*Cycloheximide Treated fibroblast sequencing QC*

The cycloheximide (CHX) cohort was sequenced at two different facilities. Initially a small number of undiagnosed patient fibroblasts were sequenced after CHX treatment (n=14) and to improve the sample size we included 65 samples from an unpublished *COL6A*-related disease cohort in our lab. We rationalized that CHX treatment would preserve aberrant splice junction reads that would otherwise be degraded by nonsense mediated decay.

To assess the impact of cycloheximide (CHX) treatment on sequencing and alignment quality, we compared the CHX-treated cohort to the untreated fibroblast cohort. Read depths were similar but the CHX cohort had higher numbers of detected genes (average 25,956 versus 20,479). To evaluate the effectiveness of CHX in inhibiting NMD, we examined splice metrics. While splice junction counts (average 73,739,045 versus 78,987,904) and annotated splice junction counts were slightly lower in the CHX cohort, the percentage of non-annotated splice junctions was higher (average 0.12% vs 0.004%). Likely, splice junctions normally degraded (such as poison exons) are detected more frequently due to inhibition of NMD. We further confirmed successful CHX treatment by examining splicing patterns in the *SRSF2* gene, which has NMD sensitive and insensitive transcripts[43]. Isoform switching to NMD sensitive transcripts in *SRSF2* is observed relative to untreated fibroblasts.

*Splice Outlier Bed File*

For FRASER, chromosomal coordinates corresponding to reported outlier regions were extracted and compiled into a bed file. For MINTIE, outlier calls were filtered to retain only splicing-related categories by excluding variants classified as "DEL," "INS," "FUS," and "IGR." The remaining data were further filtered to include only events with a FDR ≤ 0.05, and the associated chromosomal coordinates were extracted into a bed file.

For rMATS-Turbo, results were distributed across multiple output files per sample. Five MATS JC files were selected for analysis. Coordinates were extracted from each file after filtering for an FDR ≤ 0.05. For A3SS and A5SS events, the 'longExonStart_0base' and 'longExonEnd' columns were used; for MXE events, the '1stExonStart' and '2ndExonEnd' columns were used; for RI events, the 'riExonStart_0base' and 'riExonEnd' columns were used; and for SE events, the 'exonStart_0base' and 'exonEnd' columns were used.

The resulting bed files from FRASER, MINTIE, and rMATS-Turbo were concatenated into a single file. For any entries where the start and end coordinates were identical, 1 bp was added to the end coordinate to comply with bedops format requirements. The combined bed file was then sorted using bedops (v2.4.41) and intersected with the filtered variant bed file using the --range -50:50 -e set operation to identify variants overlapping splice outlier regions. The resulting overlapping variant list was further filtered based on SpliceAI score threshold greator than 0.1, and individual variants were visually inspected in IGV. For cases in which parental sequencing data were available, variants overlapping splice outliers were uploaded to seqr and evaluated for segregation using built-in search functionalities.
